# Intrahost evolution leading to distinct lineages in the upper and lower respiratory tracts during SARS-CoV-2 prolonged infection

**DOI:** 10.1101/2024.03.03.24303157

**Authors:** Majdouline El Moussaoui, Sebastien Bontems, Cecile Meex, Marie-Pierre Hayette, Marie Lejeune, Samuel L. Hong, Simon Dellicour, Michel Moutschen, Nadine Cambisano, Nathalie Renotte, Vincent Bours, Gilles Darcis, Maria Artesi, Keith Durkin

## Abstract

Accumulating evidence points to persistent severe acute respiratory syndrome coronavirus 2 (SARS-CoV-2) infections in immunocompromised individuals as a source of genetically divergent, novel lineages, generally characterised by increased transmissibility and immune escape. While intrahost evolutionary dynamics of the virus in chronically infected patients have been previously reported, existing knowledge is primarily based on samples obtained from the nasopharyngeal compartment. In this study, we investigate the intrahost evolution and genetic diversity that accumulated during a prolonged SARS-CoV-2 infection with the Omicron sublineage BF.7, estimated to have persisted for over one year in an immunosuppressed patient. Based on the sequencing of eight viral genomes collected from the patient at six time points, we identified 86 intrahost single-nucleotide variants (iSNVs), two indels, and a 362 bp deletion. Our analysis revealed distinct viral genotypes in the nasopharyngeal (NP), endotracheal aspirate (ETA), and bronchoalveolar (BAL) samples. Notably, while significant divergence was observed between NP and BAL samples, most of the iSNVs found in ETA samples were also detected in NP or BAL samples. This suggests that NP samples may not offer a comprehensive representation of the overall intrahost viral diversity. Nonsynonymous mutations were most frequent in the spike and envelope genes, along with loss-of-function mutations in ORF8, generated by a frameshift mutation and a large deletion detected in the BAL and NP samples, respectively. Using long-range PCR on SARS-CoV-2 samples sequenced as part of routine surveillance, we validated that similar deletions causing ORF8 loss of function can be carried by SARS-CoV-2 during acute infection. Our findings not only demonstrate that the Omicron sublineage BF.7 can further diverge from its already exceptionally mutated state but also highlight that patients chronically infected with SARS-CoV-2 can develop genetically specific viral populations across distinct anatomical compartments. This provides novel insights into the intricate nature of viral diversity and evolution dynamics in persistent infections.

## Introduction

The SARS-CoV-2 pandemic has been characterised by the repeated emergence of divergent lineages with a significant growth advantage over contemporary variants [1,2]. These variants of concern (VOCs) generally exhibit enhanced transmissibility and the ability to evade pre existing acquired immunity, posing significant challenges to the containment of SARS-CoV-2 spread. The World Health Organization (WHO) has designated five VOCs of SARS-CoV-2, namely, the Alpha (B.1.1.7), Beta (B.1.351), Gamma (P.1), Delta (B.1.617.2), and lastly Omicron (B.1.1.529) in November 2021 [2]. Compared with previous variants, Omicron encompassed a substantially larger number of unique mutations in the spike gene, contributing to antigenic distance and probably occurring at least partly in the context of a chronic infection. The emergence of Omicron raised immediate global concerns, driven by its rapid global spread attributed to its remarkable ability to evade immune responses induced by prior vaccination or infections, increased transmission rate, and high tropism for nasal epithelial cells [3–5]. Subsequently, several antigenically drifted sublineages of Omicron (e.g., BA.4/5, BA.2.75.2, BA.4.6, BQ.1.1, and XBB) emerged and supplanted prior subvariants [6]. Compared to the BA.2 lineage, BA.4 and BA.5 have demonstrated increased escape from antibody neutralization, attributed to mutations at antigenically potent RBD positions, notably L452R and F486V [7]. Although a large fraction of the mutations are localised in the Spike’s receptor-binding domain (RBD) or N-terminal domain (NTD), accumulating evidence suggests that mutations impacting viral genes outside of spike play critical roles in the fitness of Omicron sublineages [8]. More recently, the second half of 2023 saw the emergence of the novel lineage BA.2.86, an Omicron sublineage, which had accumulated over thirty mutations in the spike protein compared to its BA.2 progenitor. Although BA.2.86 initially showed modest growth, the acquisition of additional mutations led to the daughter lineage JN.1, which has subsequently become the dominant lineage worldwide [9].

Newly emergent VOCs generally perch atop a long ancestral phylogenetic branch, rooted in a historic lineage absent from the current variant landscape [6]. While SARS-CoV-spillover/spillback to and from an animal host could potentially generate similar long phylogenetic branches [10], recent evidence suggests that immunocompromised individuals with persistent infections are the main source of these divergent lineages [2,11]. Since the early stages of the pandemic, long-term replication has been documented in chronically infected immunocompromised individuals, accelerating intrahost viral evolution and leading to the acquisition of amino acid changes that often overlap with those identified in VOCs [12–14]. Therefore, monitoring of SARS-CoV-2 mutations and understanding intra-host viral evolution in immunocompromised individuals is a priority.

In this report, we characterised the intrahost genetic diversity and evolution of SARS-CoV-2 in an immunocompromised patient with B-cell depletion due to diffuse large B-cell lymphoma (DLBCL), showing persistent SARS-CoV-2 Omicron BF.7 replication, suspected to have spanned over a year. In contrast to most chronic SARS-CoV-2 infections reported in the literature, we sequenced SARS-CoV-2 from different specimen sites collected on the same day, including nasopharyngeal swab (NP), endotracheal aspirate (ETA), and bronchoalveolar lavage (BAL) samples. The resulting sequences revealed a deep divergence in the viral lineages at opposite ends of the respiratory tract, with a mixture of viral populations in the ETA sample. These findings highlight that NP samples only capture a subset of the diversity present in chronic infection and that a negative test does not rule out the presence of a SARS-CoV-2 reservoir in immunocompromised individuals.

## Results

### The identification of a persistent SARS-CoV-2 infection in a immunosuppressed host

The patient, in their sixties, suffered from follicular lymphoma (Grade 2, clinical stage IV), which subsequently progressed into diffuse large B-cell lymphoma (DLBCL). During the end of 2021, they underwent treatment with four cycles of rituximab with cyclophosphamide, doxorubicin, vincristine and prednisone (R-CHOP), followed by two cycles of rituximab alone, resulting in a complete remission of the disease. Since February 2022, the patient has been receiving bimonthly infusions of rituximab as maintenance therapy. In addition, the patient’s medical history was also notable for a first mild episode of COVID-19 in January 2022, which resolved over a few days. In April, reverse-transcriptase polymerase chain reaction (RT-PCR) for SARS-CoV-2 on NP was negative. Of note, the patient had received three doses of vaccine (two doses of ChAdOx1-S followed by one of BNT162b2) between March and October 2021, resulting in detectable anti-trimeric spike protein-specific IgG antibodies in June 2021.

In July 2022, they were admitted for fever, cough, shortness of breath, haemoptysis, diarrhoea, and generalised weakness, evolving over a period of four days. Chest computed tomography (CT) showed diffuse and bilateral ground-glass opacities with moderate involvement (20 to 25%) taking consolidated appearance in the middle lobe, as well as a small bilateral pleural effusion. SARS-CoV-2 PCR performed on an NP swab was positive (RdRp Gene, cycle threshold [Ct] = 35), and the patient was diagnosed with COVID-19 pneumopathy. They were treated with a five-day course of Molnupiravir and required transient oxygen supplementation, but eventually evolved favourably and were discharged home seven days after admission. The maintenance therapy with Rituximab was also discontinued.

During the subsequent months, the patient reported intermittent fatigue, low-grade fever, cough, blood-tinged sputum, and dyspnea. Chest CT revealed diffuse ground-glass opacities with new upper left and middle lobe consolidations suggestive of organising pneumonia (OP). After exclusion of infectious alternative diagnoses including notably a negative RT-PCR for SARS-CoV-2 on NP sample, treatment with methylprednisolone (8 mg/day) was initiated in September 2022, resulting in partial resolution of the symptomatology over a period of three weeks. However, shortly after discontinuing corticosteroids, symptoms recurred, prompting their reintroduction by the end of October 2022. Between December 2022 and April 2023, the clinical condition was marked by systematic recurrence of respiratory symptoms following each attempt of corticosteroid discontinuation. Of note, RT-PCR for SARS-CoV-2 performed on BAL sample remained negative in April 2023.

In May 2023, the patient was hospitalised for fever, cough, and moderate respiratory failure that required oxygen supplementation. Chest CT showed a marked increase in diffuse and bilateral ground-glass opacities with severe lung involvement (75%). SARS-CoV-2 RT-PCR on NP swab was positive, confirming the diagnosis of COVID-19 pneumopathy. The patient was treated with dexamethasone and a seven-day course of antibiotic therapy due to a high suspicion of bacterial co-infection. Following an initial clinical amelioration, permitting discharge home with oxygen supplementation and oral corticosteroids, they were readmitted in June due to a subsequent exacerbation of respiratory insufficiency. This progression necessitated an eight-day stay in the intensive care unit (ICU) for the administration of high-flow nasal cannula therapy. After multiple rounds of antibiotic therapy for bacterial superinfections, intravenous immunoglobulin, and high-dose corticosteroid pulses, they gradually improved and were able to return home after approximately three months of hospitalisation. The persistence of the infection during the period was confirmed by repeated positive RT-PCR tests on NP, AET, and BAL samples, with Ct values ranging from 16.1 to 26.4, between May 2023 and September 2023 (126 days). Finally, in October 2023, the RT-PCR test yielded a negative result. The clinical course of the infection is summarised in Figure 1, while the evolution of imaging is displayed in Supplementary Figure S1.

**Figure 1.**
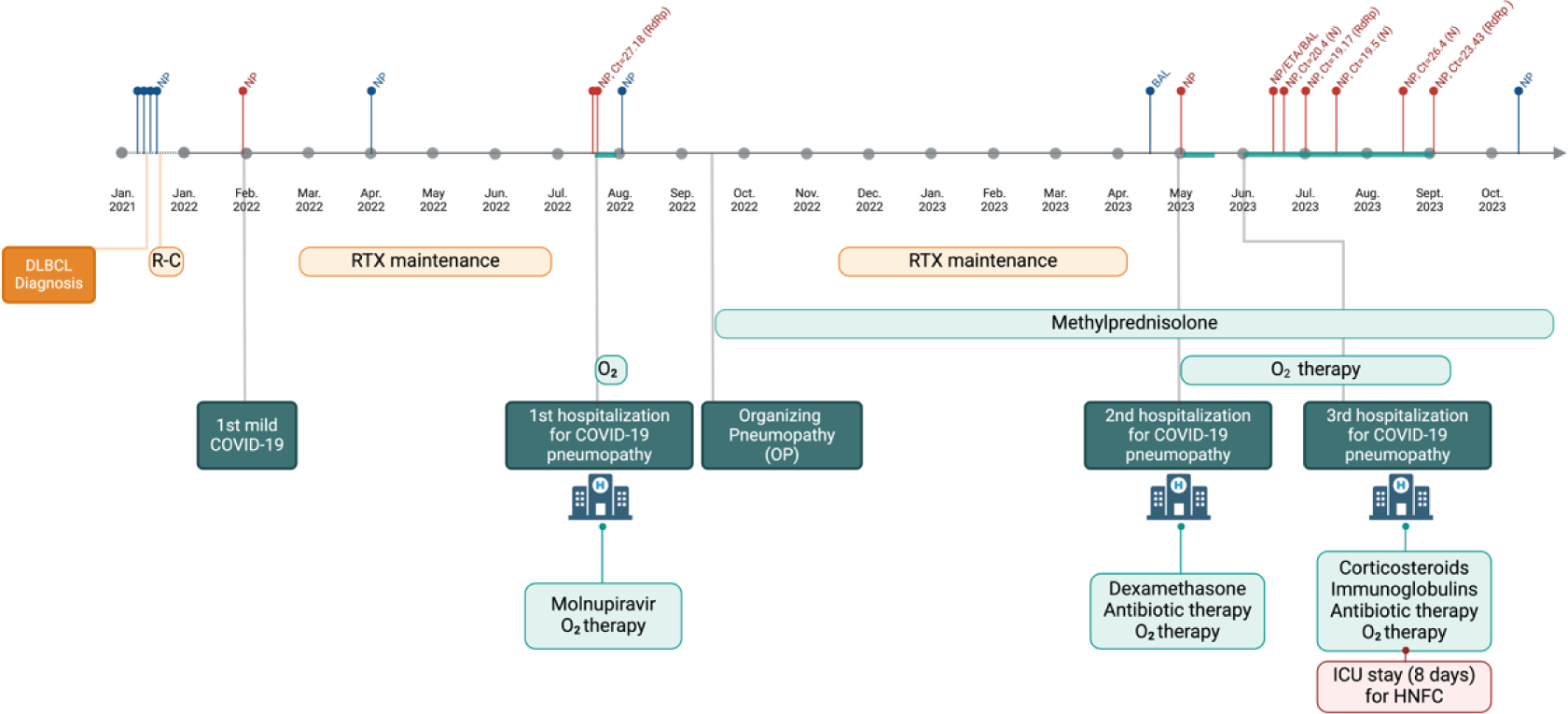
Timeline of the clinical history and disease course. The figure illustrates the timeline progression of the patient’s COVID-19 infection along with relevant clinical events. The top section displays the SARS-CoV-2 RT-PCR results and Ct values in NP, ETA, and BAL samples over time. Positive test results are indicated in red, while negative tests are represented in blue. The lower section provides a comprehensive overview of the patient’s clinical history including the history of diffuse large B-cell lymphoma (DLBCL) and its treatment, three hospitalizations (July 2022, May 2023, and from June 2023 to September 2023, including an eight-day ICU stay in June 2023) related to COVID-19, and the therapeutic interventions administered to the patient. Abbreviations: BAL = broncho-alveolar lavage, Ct = cycle threshold, DLBCL = diffuse large B-cell lymphoma, ETA = endotracheal aspirate, HFNC = High-flow nasal cannula therapy, ICU = intensive care unit, NP= nasopharyngeal swab, R-C = Rituximab with cyclophosphamide, doxorubicin, vincristine and prednisone (R-CHOP), RTX = Rituximab.

### Genomic sequencing points to chronic infection lasting over a year

In total, we sequenced eight samples from the patient collected over an 85-day period, between June and September 2023. These samples were found to be derived from the almost extinct Omicron BA.5 sublineage BF.7 (Fig. 2A). This sublineage saw its Belgian peak in October 2022, accounting for 17.4% of sequenced samples (Fig. 2B). The sublineage declined in the following months, with the last Belgian example sequenced in February 2023. The patient’s consensus genomes were placed onto the existing SARS-CoV-2 phylogenetic tree using the UShER tool [15]. They sat at the end of a long phylogenetic branch, separated by over 25 single nucleotide variants (SNVs) from their closest relatives, which were collected towards the end of 2022 (Fig. 2C).

**Figure 2.**
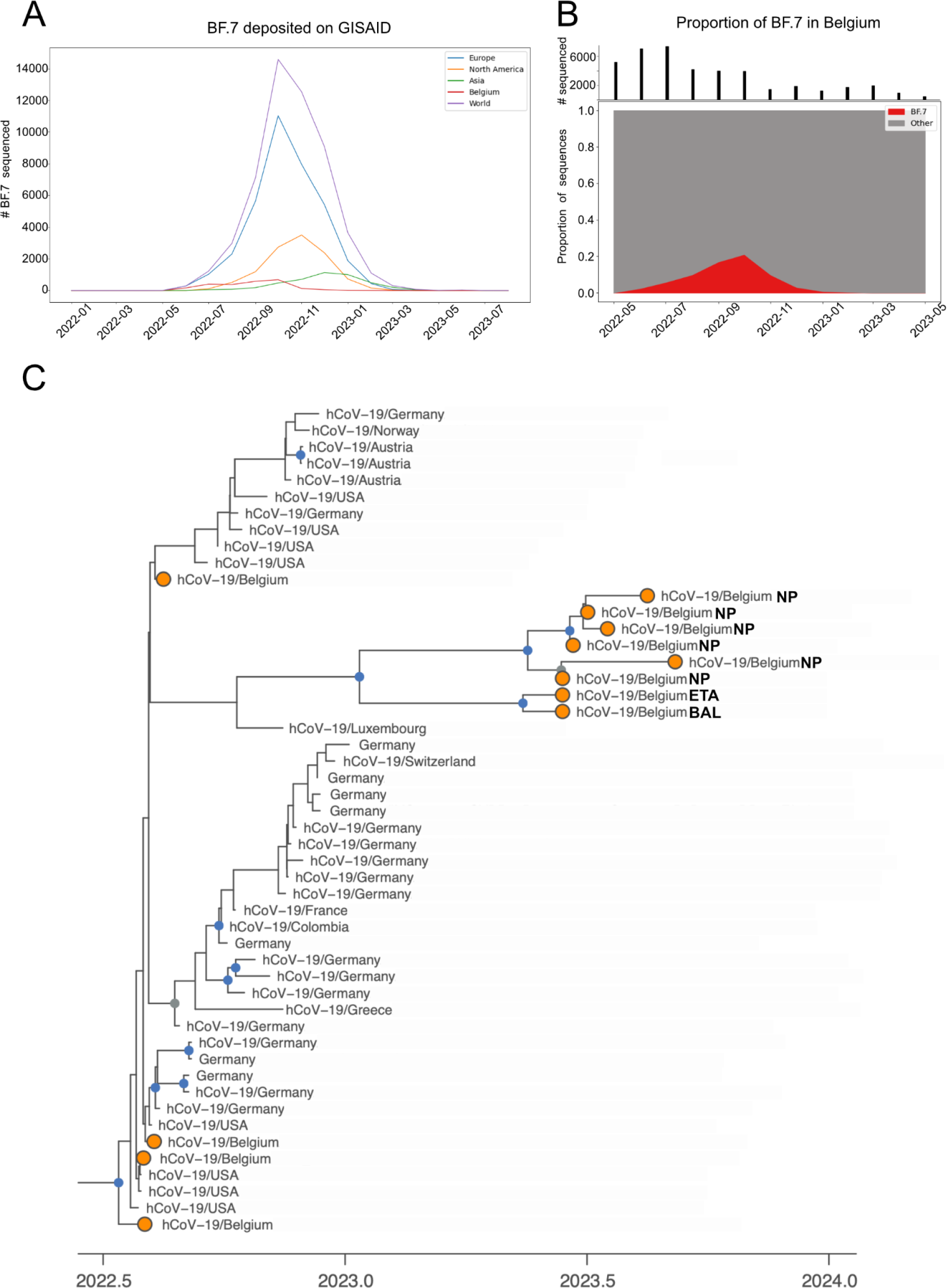
Dynamics of BF.7 sublineage and phylogenetic tree. (A) Temporal distribution of BF.7 sublineage (BA.5.2.1.7) samples from GISAID. (B) Proportion of public Belgian genomes from GISAID belonging to BF.7 sublineage sampled between May 2022 and May 2023. The number of samples that were sequenced each month is indicated above. (C) Maximum clade credibility (MCC) tree resulting from a time-scaled phylogenetic inference carried out to display the evolutionary relationships among the eight SARS-CoV-2 BF.7 sequences from the patient (NP for nasopharyngeal, ETA for endotracheal aspirate, and BAL for bronchoalveolar lavage) and the most closely related genomes identified in the UShER global phylogenetic tree. Belgian genomes are marked in orange, and nodes with a posterior probability above 0.7 and 0.9 are coloured in grey and blue, respectively.

The patient tested positive for SARS-CoV-2 in January 2022 with a subsequent negative test in April. At this time, BF.7 was observed at very low frequencies worldwide (Fig. 2A) and was not detected in Belgium until May 2022 (Fig. 2B), making this first infection an unlikely time point for the establishment of a persistent infection. The patient again tested positive in July 2022, and at this time, BF.7 was the fifth most common lineage observed in Liège, representing 6.3% of samples sequenced during the week they tested positive (Supplementary Fig. S2). The patient had negative tests in August 2022 and in April 2023, but again tested positive in May 2023 and was consistently positive in subsequent tests (Fig. 1). Unfortunately, only samples from June 2023 forward were sequenced, and as a consequence, it is impossible to draw a definitive conclusion. Nevertheless, based on the viral lineage observed in the samples taken from June 2023 on, it would appear that the infection acquired in July 2022 never cleared (despite intervening negative PCR tests) and suggests the patient was chronically infected for over a year.

### Distinct viral populations in the upper and lower respiratory tracts

While the presence of a long branch leading to the patient samples is expected in a prolonged infection, the relative placement of the consensus genomes was initially somewhat surprising. Notably, the six NP samples formed a cohesive cluster, whereas the BAL and ETA samples occupied a distinct branch that showed early divergence from the NP samples, indicating a long period of independent evolution. Across all samples, we identified eighty-six unique intrahost single-nucleotide variants (iSNVs; Fig. 3A), two indels and a 362 bp deletion (MN908947.3:27818-28180) distributed in accessory protein-coding genes ORF7a and ORF8 (Fig. 3B, Supplementary Fig. S4, Table S1). All iSNVs had an allele frequency of 18% or more in at least one sample. Interestingly, substantial disparities were observed in genetic variants found in upper and lower respiratory tract samples. In the first NP sample taken in June, 2023, we observed 29 iSNVs as well as the 362 bp deletion, all with an allele frequency of 87% or higher (Fig. 3A). In contrast, in the BAL sample collected on the same date, 54 iSNVs and two indels were found, with frequencies varying between 19 and 99% (Fig 3A). Remarkably, only four of these genetic variations were found in both samples (Fig. 3B), explaining the significant divergence observed in the branches of the phylogenetic tree (Fig. 1C). Regarding the ETA sample, we observed 68 iSNVs with allele frequencies ranging from 9 to 97%. Most of iSNVs found in ETA samples were also identified in NP or BAL samples. Only two iSNVs were unique to the ETA sample (Fig 3B). In comparison to the BAL and ETA samples, iSNVs identified in NP samples generally showed high allele frequencies, with variants appearing and disappearing relatively abruptly between samples (Fig. 3A). Over an 85-day period, we observed the emergence of six iSNVs and one reversion in the subsequent five NP samples. Notably, in the last sample, only three of the new iSNVs and the reversion persisted, as three had been lost in the intervening time (Fig. 3A).

**Figure 3.**
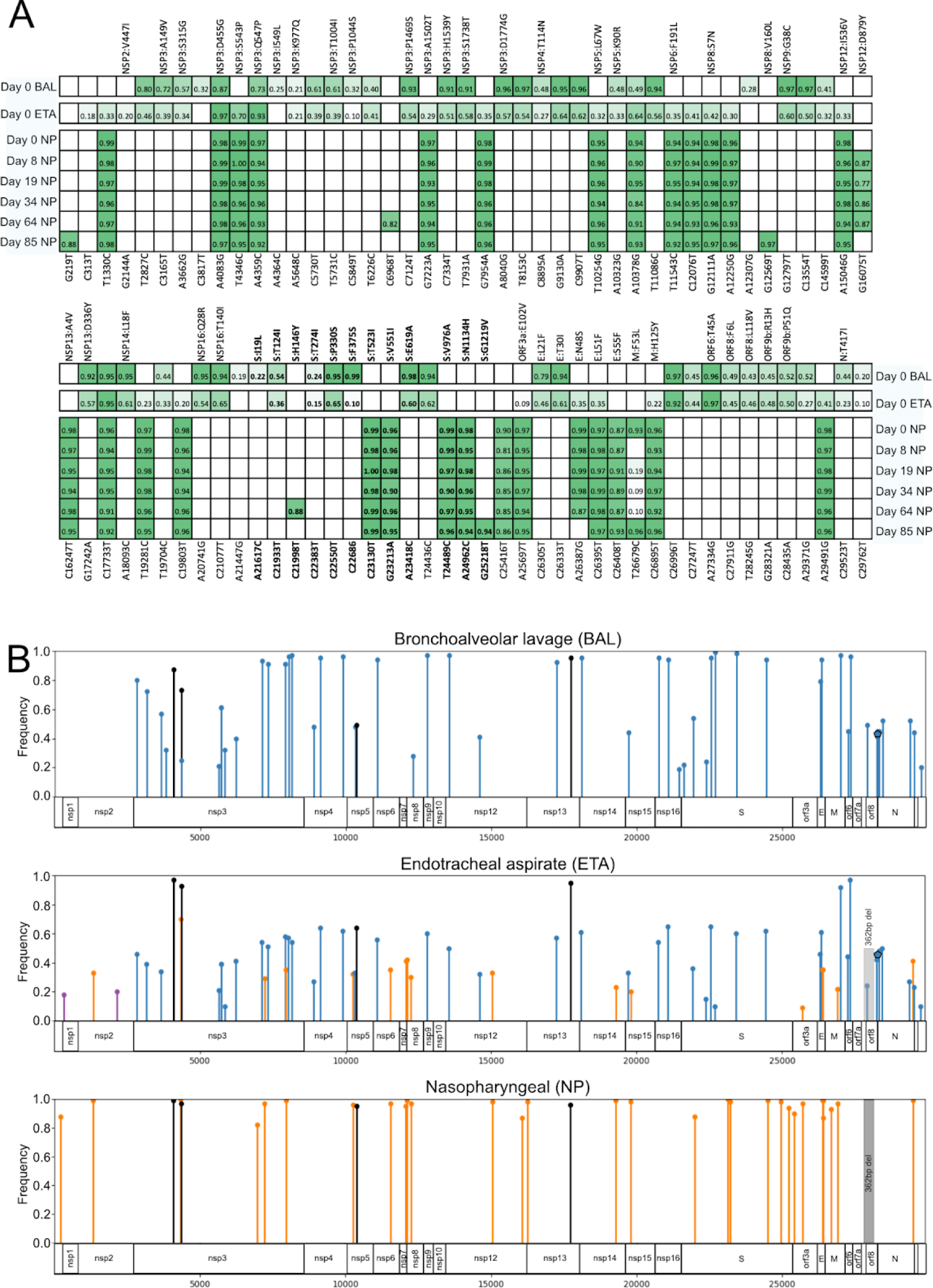
Evolution of patient-specific variants overtime according to different anatomical compartments. (A) Frequency of iSNV across the upper and lower respiratory tracts over time, day 0 corresponds to the first samples sequenced, taken June 2023. The top panel shows iSNV at the 5’ end of the virus; the bottom shows the 3’ end. Genomic position corresponding to ancestral Wuhan-1 (MN908947.3) is shown on the bottom, while amino acid changes are indicated on top. Colour intensity reflects allele frequency. Abbreviations: NP = nasopharyngeal, BAL = bronchoalveolar lavage, ETA = endotracheal aspirate. (B) Patient-specific mutations from upper and lower respiratory tracts are mapped to the SARS-CoV-2 genome, displaying viral proteins. The graph shows the frequency of alleles for variants found in NP, BAL, and ETA samples collected at six different time points. The height of the line represents the allele frequency. The highest allele frequency observed is used for NP samples at the six time points combined. Variants that are exclusively found in BAL and ETA samples are represented in blue, those found only in NP and ETA are shown in orange, and ETA-exclusive variants are shown in purple. Variants that are present in all the samples are represented in black. iSNVs are represented by circles, indels by polygons, and deletions by shaded rectangles.

### iSNVs mutational patterns

The iSNV mutational landscape was dominated by C-U transitions (Fig. 4A), which is in line with previous reports [16–18]. Despite treatment with Molnupiravir [19] the frequency of G-to-A iSNV was similar to the G-to-A SNVs seen in ancestral BF.7 (Supplementary Fig. S4). When we compared the mutation patterns between samples collected at different anatomical sites, we did not observe a marked difference between BAL and NP samples (Fig. 4A). Additionally, the frequency of synonymous and nonsynonymous mutations in iSNVs was similar in BAL and NP samples (Fig. 4B). Considering all iSNVs, we further examined the proportion and number per kilobase of synonymous versus nonsynonymous changes in SARS-CoV-2 proteins impacted by at least three iSNVs. The spike and envelope proteins showed the highest frequency of nonsynonymous changes (Fig. 4C-D). We then investigated NP and BAL-specific iSNVs for the three SARS-CoV-2 proteins with the highest number of variants. We observed a similar pattern for spike and envelope in both BAL and NP samples. For NSP3 in the NP samples there was no difference in the frequency or the number of synonymous and nonsynonymous changes, while in the BAL sample there was a modest difference (Fig. 4E-F). Deep mutational scanning of the complete XBB.1.5 spike reported by Dadonaite and colleagues [20] suggests that key changes in the BAL spike include the F375S and E619A, both of which are predicted to increase ACE2 binding [20]. Conversely, the substitution T523I in NP samples was predicted to enhance ACE2 binding, while the substitution V976A may contribute to immune escape, although at the expense of ACE2 binding [20] (Supplementary Table S2).

**Figure 4.**
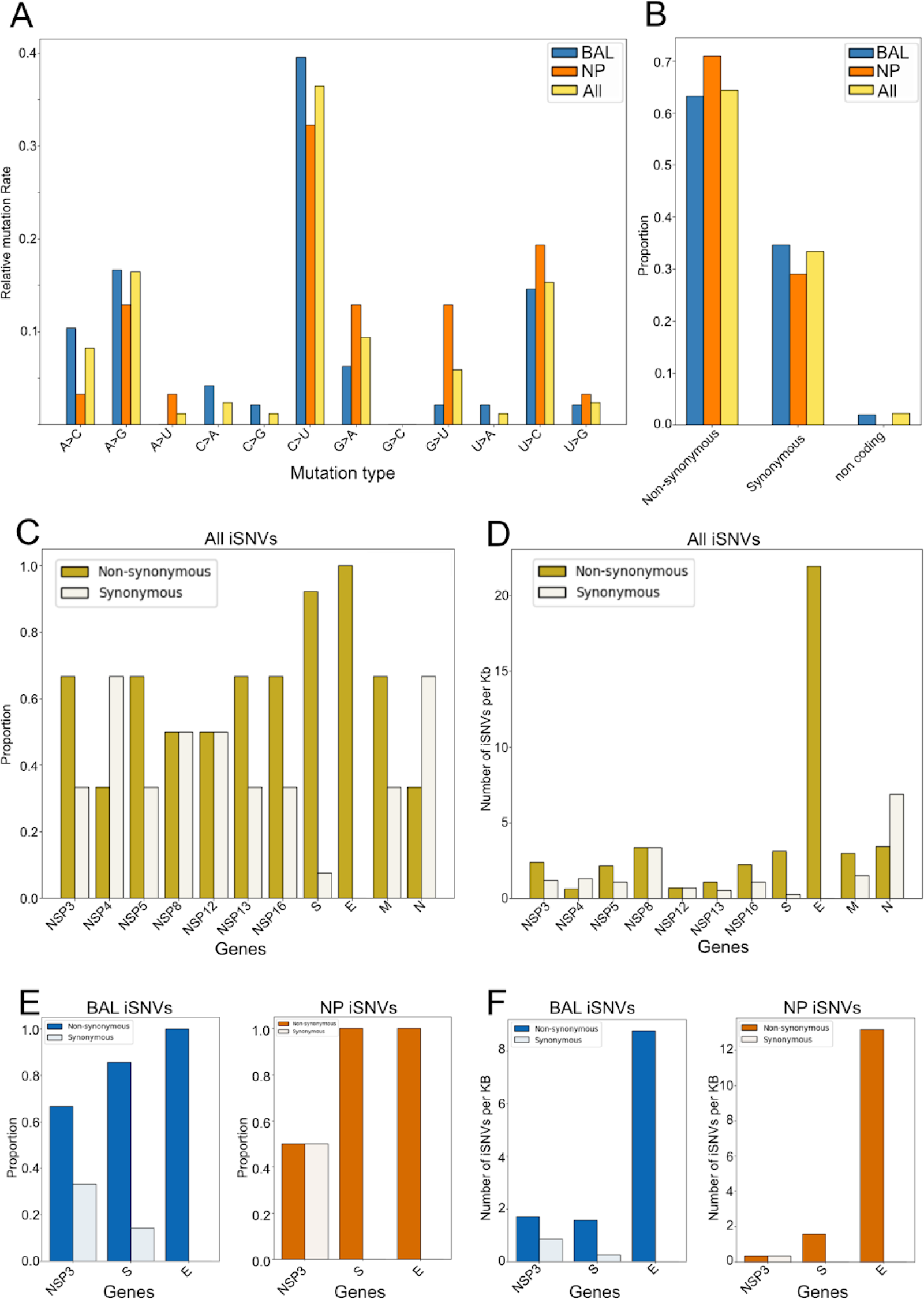
Intrahost variant impacts. (A) Mutation spectra for all iSNVs observed; (B) Frequency of synonymous versus nonsynonymous mutations in BAL iSNV, NP iSNV and all iSNV combined; (C) Frequency of synonymous versus nonsynonymous mutations specifically in viral proteins with three or more iSNVs; (D) Quantification of iSNV per kb in viral proteins with three or more iSNV; (E) Frequency of synonymous versus nonsynonymous mutations in BAL and NP for three viral proteins (NPS3, spike and envelope); (F) Frequency of synonymous versus nonsynonymous mutations in BAL and NP for three viral proteins (NPS3, spike and envelope) with a presentation of iSNVs per kilobase.

### ORF8 knockouts were also observed in acute infections

The frameshift identified in the BAL sample and the large deletion detected in the NP sample resulted in loss of function (LOF) mutations within ORF8. During the routine genomic surveillance of SARS-CoV-2, we have noticed large putative deletions in the ORF7a/b/8 region (discerned through a lack of coverage in the region and, in some instances, via reads spanning the breakpoint). To confirm that such deletions can be found in acute infections, we performed long range PCR on two routine samples suspected to have these deletions. The results revealed an XBB.1.5 sublineage carrying a 306 pb deletion within ORF8 and a CH.1.1 sublineage carrying an 872 bp deletion, which removes ORF7a, ORF7b and ORF8 (Supplementary Fig. S5). We used UShER to place these genomes on the SARS-CoV-2 phylogenetic tree and found that most of the adjacent genomes had either a gap or Ns in the same region (Supplementary Table S6), strongly indicating the presence of the respective deletions.

## Discussion

The case reported above documents a persistent SARS-CoV-2 infection in a patient undergoing B-cell depletion therapy with anti-CD20 therapy for diffuse large B-cell lymphoma. The chronic infection was confirmed through migratory lung infiltrates and positive RT-PCR tests from samples collected in the upper and lower respiratory tracts. Determining the exact duration of chronic infection in this patient is a challenge, as sequencing of samples only began in June 2023, and the patient intermittently tested negative for SARS-CoV-2. Nonetheless, we could confirm via sequencing that the infection endured for more than three months (between June and September 2023) and, based on the archaic lineage observed in the viral sequences, it is likely that the infection was acquired in June 2022. These findings suggest a prolonged infection lasting over a year and highlight the difficulty in diagnosing a low-grade SARS-CoV-2 viral infection in an immunocompromised patient. One of the strengths of this study is the examination of multiple samples from different anatomical compartments, and sampling at various time points, providing a broader understanding of the infection. A chronic infection could potentially be missed if data were only based on samples collected from the same site and over shorter periods. The extended reconstitution period of the B-cell population after rituximab treatment (six to twelve months), leading to significantly impaired autologous B-cell function stands out as one of the most influential risk factors for prolonged SARS-CoV-2 infection and viral intra-host evolution among immunocompromised individuals [21,22]. Consequently, this population represents a crucial target for genetic surveillance of viral evolution in immunocompromised patients.

In this study, we obtained sequence data from eight positive specimens, covering six time points, including six nasopharyngeal samples, one endotracheal, and one bronchoalveolar sample. The mutational pattern showed predominance of C-U transitions, which is in line with the pattern of substitutions commonly observed in SARS-CoV-2 [16,23]. While the patient received Molnupiravir before sample collection, there was no characteristic excess of G-to-A mutations observed in the iSNV [19], suggesting this treatment did not have a major impact on the evolution of the viral population in the patient. Furthermore, we observed a higher proportion of non-synonymous changes relative to synonymous changes in the spike and envelope genes, with no discernible difference between NP and BAL specimens.

The interpretation of amino acid changes is commonly based on their deviation from the Wuhan-Hu-1 reference sequence. However, following ∼4 years of selection and drift the lineages that now dominate differ from the reference genome at a number of positions across the viral genome, especially in regions coding for viral proteins that are targeted by the host adaptive immune system. Like Omicron lineages BA.2 and BA.5, the BF.7 sublineage carries the amino acid change S375F in the spike RBD. While the S375F mutation was still present in NP samples, the sequence obtained from the BAL sample reverted to the wild-type sequence. Recently deep mutational scanning using the XBB.1.5 and BA.2 spikes by Dadonaite and colleagues in 2023 [20] showed that a number of amino acid changes at position 375 (including the reversion to the reference) can increase ACE2 binding and highlights that the impact of an amino acid change is often context dependent. Among the remaining 11 amino acid changes observed in spike, E619A (BAL) and T523I (NP) are predicted to increase ACE2 binding, while V976A (NP) facilitates immune escape but reduces ACE2 binding (Supplementary Table S2). S:P330S (BAL) is predicted to cause a modest increase in ACE2 binding and has been observed in other chronic infections involving immunosuppressed individuals [14,20,24]. All the spike amino acid changes, except for the revertant at position 375, are currently observed at a low frequency in GISAID (Supplementary Table 2). However, if we use persistent infections involving pre-VOC lineages as a guide for Omicron persistent infections, it is reasonable to assume that some of the amino acid changes observed in the current case may appear in future lineages that rise to prominence, especially if these lineages are derived from a BA.5/4 ancestor.

The spike protein plays a central role in cellular entry and represents a primary target for the adaptive immune system. Consequently, this protein has been subject to extensive scrutiny relative to the other 14 open reading frames of SARS-CoV-2. However, there is clear evidence that genetic variations outside the spike protein play a role in the pathogenicity and fitness of SARS-CoV-2. Recent work using a recombinant SARS-CoV-2 consisting of the spike of Omicron (BA.1) and the backbone of an ancestral isolate showed higher virulence than BA.1, with mutations in nsp6 identified as the main driver of this difference in phenotype [25]. It has also been noted that BA.5 outcompeted BA.4 despite having identical spike proteins [26]. Among the amino acid changes observed outside the spike in the current case, a number have previously been associated with chronic infections. Notably, E:T30I has been observed in multiple chronic infections [26,27] but is rarely seen in circulating isolates (0.01% of GISAID genomes in January 2024) and appears to have a negative impact on the virus’s ability to transmit to new hosts [27,28]. Likewise, H125Y in the M protein, has also been identified in multiple chronic infections [24] and has been observed to arise in cell culture settings [29]. It is infrequent in sequenced genomes (0.1% in GISAID as of January 2024), yet computational analyses suggest a fitness advantage for the virus harbouring this change [24,28]. Unlike the previous amino acid changes, NSP3:K977Q has previously been observed in a VOC (Gamma, P.1) and has been detected in the context of chronic infections as well as within cryptic lineages sampled from wastewater [30–32]. This amino acid change enhances the activity of the SARS-CoV-2 papain-like protease, potentially modulating the host’s innate immune response [33]. Finally while NSP5:K90R has not been implicated in chronic infections, of the amino acid changes observed in the patient it was the one with the highest impact on fitness based on the work of Bloom et al [28] (Supplementary Table 1) and like NSP3:K977Q it may play a role in modulating the innate immune response [34]. These findings underscore the diverse impacts of non-spike protein changes on viral transmission dynamics and host immune system modulation.

Beyond single amino acid changes, sequencing of NP and BAL samples revealed a large deletion and a frameshift mutation in the ORF8 region, resulting in a complete loss of function of the viral accessory protein. Notably, ORF8 is recognized as one of the most hypervariable regions in the SARS-CoV-2 genome and presents the highest mutation density among non-structural proteins [28]. Since the early stages of the COVID-19 pandemic, various large deletions affecting ORF8 have been documented, with some found to reduce COVID-19 severity [35–39]. The mutational landscape of ORF8 over the past ∼four years of the pandemic suggests a strong selective pressure driving rapid evolution, leading to ORF8 knockouts across multiple SARS-CoV-2 lineages [35,36,39,40]. Several studies have reported evidence of relaxed purifying selection acting on ORF8, along with other accessory proteins [28], with ORF8 knockout clusters exhibiting a growth rate advantage [41]. Recent research by Kim and colleagues has proposed a biological mechanism underlying positive selection for ORF8 knockouts [42]. They demonstrated that ORF8 can downregulate virion spike protein levels, thereby facilitating immune evasion by reducing the presence of spike on the surface of infected cells [42]. Conversely, the absence of ORF8 increases spike protein levels on the virion, potentially enhancing its ability to infect host cells. This dynamic may confer a net benefit depending on the context. In immunocompromised individuals, viral strains lacking ORF8 could have “the best of both worlds” by exhibiting heightened infectivity without eliciting an immune response against infected cells due to reduced Spike protein exposure on the cell membrane. Moreover, viral strains lacking a complete ORF8 have been reported in immunocompromised patients with chronic SARS-CoV-2 infections, underscoring the importance of detecting ORF8 knockouts in SARS-CoV-2 genomic surveillance efforts [43].

Our phylogenetic analysis revealed that NP samples formed a cohesive cluster, while BAL and ETA samples occupied a branch that diverged from NP samples early on, indicating a prolonged period of independent evolution. Notably, in the initial samples collected on July 14, 2023, the BAL sample carried twenty-four iSNVs (allele frequency >70%) not seen in the NP sample. The NP sample carried twenty-six patient-specific variants (allele frequency >85%) that were not observed in the BAL sample. For reference, the original VOC Alpha had only twenty-three nucleotide changes compared to its ancestral lineage [44]. Although significant divergence was observed between variants detected in NP and BAL samples, most of the iSNVs found in the ETA sample were also identified in NP or BAL samples. This divergence aligns with documented viral diversity in different anatomical sites during acute SARS-CoV-2 infection, particularly when saliva and nasopharyngeal swab samples are tested concurrently [45]. Furthermore, our findings resonate with previous evidence of genetic compartmentalization in SARS-CoV-2 between pulmonary and extra-pulmonary tissues, as observed in post-mortem samples from immunocompromised patients and unvaccinated patients, both infected with SARS-CoV-2 in the first year of the pandemic [46,47].

The results presented above add to an accumulating body of evidence suggesting that in a small number of cases SARS-CoV-2 can form a reservoir in different anatomical compartments [48–50] and that in many cases this reservoir is not always detectable via conventional SARS-CoV-2 testing [48]. This highlights that further investigation on how these tissue reservoirs contribute to SARS-CoV-2 persistence is needed. These observations also have implications for the care of immunocompromised individuals as a negative SARS-CoV-2 test is not a guarantee that an infection has cleared. Alternative specimen types, such as stool should be considered [50] and any SARS-CoV-2 positive samples from immunocompromised patients should be sequenced to distinguish between acute and long-term SARS-CoV-2 infections.

## Materials and Methods

### SARS-CoV-2 genomic surveillance at the CHU of Liège

Since the beginning of the pandemic the Department of Human genetics in collaboration with the Department of Clinical Microbiology of the CHU of Liège (Belgium) has been following the evolution of SARS-CoV-2 lineages circulating the region [51–53]. We have been a member of the National Genomic Surveillance Platform for SARS-CoV-2 in Belgium since its inception in 2021 [54]. The samples used in the current study were initially collected and sequences as a part of the Belgian SARS-CoV-2 genomic surveillance effort.

### Real-time PCR and RNA extraction

In most cases SARS-CoV-2 was detected in patient samples via the cobas 6800 platform (Roche) using the cobas SARS-CoV-2 real-time PCR assay which detects the ORF1ab and E genes. Alternatively the Alinity SARS-CoV-2 assay (Abbott) or Genexpert SARS-CoV-2 assay (Cepheid) were used. Positive RNA was extracted from the nasopharyngeal swabs or bronchoalveolar lavage (300 µl) with the Maxwell 48 device and the Maxwell RSC Viral TNA kit (Promega) following the manufacturer’s instructions.

### SARS-CoV-2 whole genome sequencing

Samples with a cycle threshold (Ct) ≤ 30 on the cobas system were used for sequencing. Reverse Transcription utilised 3.3 µl of eluted RNA, 1.2 µl of SuperScript IV VILOTM Master Mix and 1.5 µl of H2O for a total volume of 6 µl. The mix was incubated at 25°C for 10 min, 50°C for 10 minutes and 85°C for 5 minutes. PCR was carried out using Q5® High-Fidelity DNA Polymerase (NEB) and the ARTIC v5.2.0 1,200 bp amplicon primer pools (https://github.com/quick-lab/SARS-CoV-2/tree/main/1200/v5.2.0_1200) PCR conditions were set up according to the recommendations of the ARTIC Network sequencing protocol (https://www.protocols.io/view/ncov-2019-sequencing-protocol-v3-locost-bp2l6n26rgqe/v3) The resultant PCR products were indexed via the Nanopore Rapid Barcoding Kit 96 V14 (SQK-RBK114.96) in the same manner described in Freed et al. 2020 [55]. Sequencing was carried out on a GridION using 10.4.1 flow cells, with Super-accurate basecalling, carried out via Guppy 6.5.7.

### Long range PCR and sequencing

In the samples carrying a deletion in the ORF7/8 region we carried out long range PCR with the primers 27_LEFT (5’-TGGATCACCGGTGGAATTGCTA-3’) and 28_RIGHT (5’-GTTTGGCCTTGTTGTTGTTGGC-3’) from Freed et al. 2020 [55]. Reverse Transcription and PCR conditions were the same as those used for ARTIC whole genome amplicon sequencing protocol. For these PCR products to retain the full-length amplicon we used the Native Barcoding Kit 96 V14 (SQK-NBD114.96) which ligates barcodes to the ends of the amplicon. Sequencing was also carried out on a GridION using 10.4.1 flow cells, with Super-accurate basecalling carried out via Guppy 6.5.7.

### Variant calling and consensus genome generation

Consensus genomes were generated via the ARTIC-network field bioinformatics pipeline (https://github.com/artic-network/fieldbioinformatics). Consensus sequences were only generated for regions with coverage of ≥30. Low frequency variants were called with LoFreq [56]. Variants were also called with Clair3 [57]. VCFs and BAMs were checked in IGV [58]. Consensus sequence for the long range PCR products were generated via bcftools consensus command (https://samtools.github.io/bcftools/bcftools.html) and manually incorporated to artic-network field bioinformatics pipeline consensus.

### Phylogenetics

Consensus genomes were placed on the existing phylogenetic tree via UShER using the phylogenetic tree generated with genomes from GISAID, GeneBank, COG-UK and CNCB. In the case of the BF.7 samples, we used the genomes identified by UShER to infer a time-scaled phylogenetic tree using the software package BEAST v1.10.5 [59] with the following settings: a skygrid demographic model, a relaxed molecular clock with a normal (8E-4,1E-4) prior distribution, and a HKY+G4+I substitution model [60,61]. Three independent Markov Chain Monte Carlo runs were performed, each running for 10^6 iterations and sampling every 10^4 states. Convergence and mixing of all parameters were assessed using Tracer v1.7 [62] and a maximum clade credibility tree was obtained using TreeAnnotator v.1.10.5 [59] after combining all results using LogCombiner v.1.10.5 [59] and discarding the first 10% of samples as burn-in.

## Data availability

The consensus SARS-CoV-2 genomes and raw reads are available on GISAID (www.gisaid.org) under accession numbers EPI_ISL_17960749, EPI_ISL_18515316, EPI_ISL_17960747, EPI_ISL_18059076, EPI_ISL_18059074, EPI_ISL_18059075, EPI_ISL_18136392, EPI_ISL_18274346. All GISAID genomes used in the analysis can be found via EPI_SET ID: EPI_SET_240229zf, doi:10.55876/gis8.240229zf. Raw reads for each sample have also been deposited on the European Nucleotide Archive (ENA) under the accession number PRJEB72450.

## Supplementary data

Sup_Table_1_intra_host_variants.xlsx is available online.

## Ethics statement

Methods of collection, testing of biological specimens and protected health information used in this study were approved by the University Hospital of Liège Ethics Review Board, Reference number: 2020-139, informed consent was received from the patient.

## Acknowledgements

Figure 1 and Supplementary Figure S1 were performed with www.biorender.com.

## Funding

This work was supported in part by the Région Wallone project WALGEMED (convention no. 1710180) and the FNRS (H.C.008.20). Sequencing was done as a part of the National Genomic Surveillance Platform for SARS-CoV-2 in Belgium. SD acknowledges support from the *Fonds National de la Recherche Scientifique* (F.R.S.-FNRS, Belgium; grant n°F.4515.22), from the Research Foundation — Flanders (*Fonds voor Wetenschappelijk Onderzoek — Vlaanderen*, FWO, Belgium; grant n°G098321N), and from the European Union Horizon 2020 projects MOOD (grant agreement n°874850) and LEAPS (grant agreement n°101094685).

## Conflict of interest

We declare no conflicts of interest.

---

**Supplementary Figure S1.**
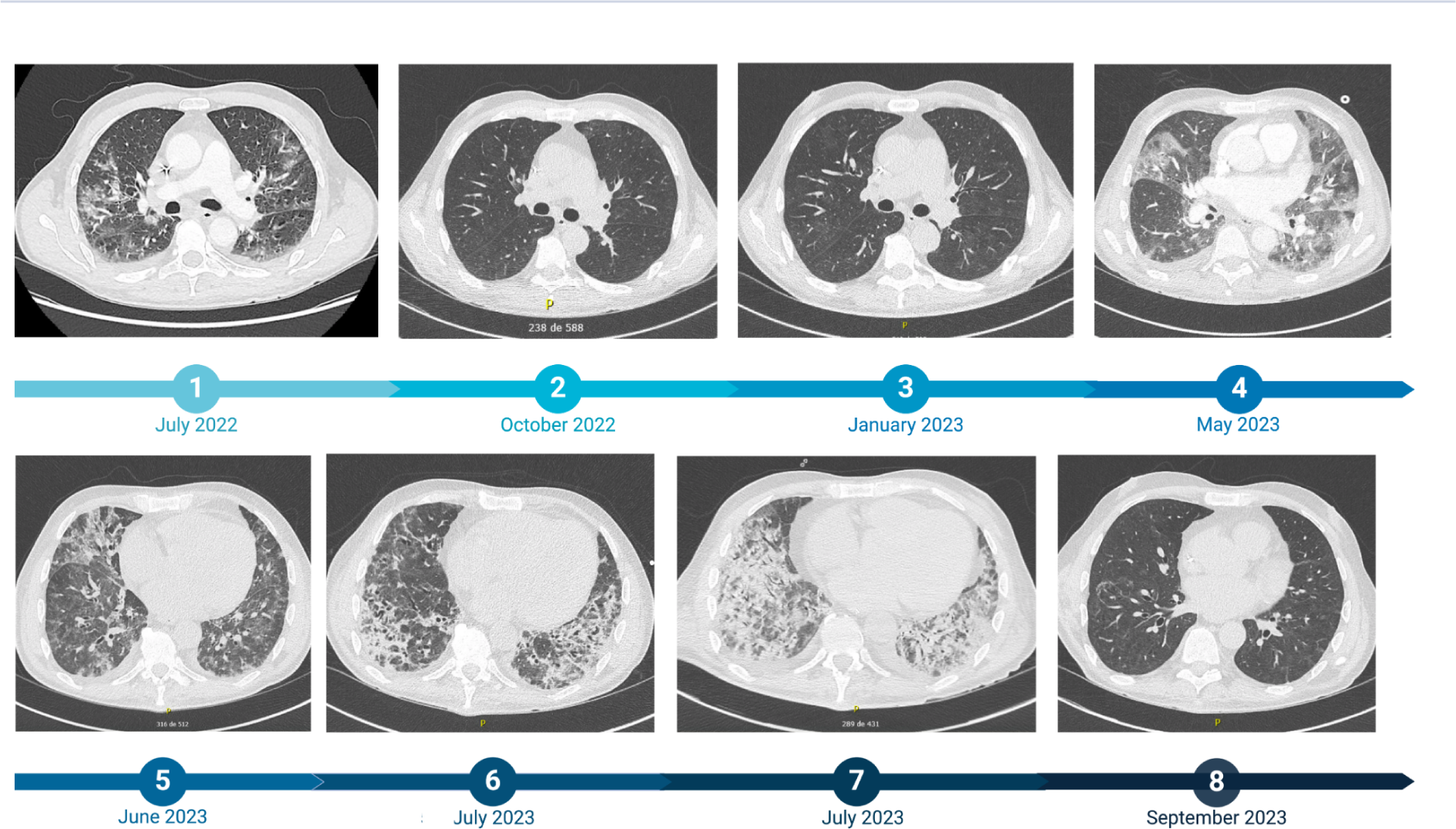
Evolution of the lesions visualized with chest CT scan over a period of one year. 1, Contrast-enhanced axial chest CT images performed during the first hospitalization showed bilateral and diffuse ground-glass opacities, consistent with moderate COVID-19 pneumonia (involvement of 20 to 25% of lung volume), and bilateral pleural effusions; 2, Axial chest CT images performed three months later showed a decrease in the extent and density of bilateral ground-glass opacities, which persist in peripheral and subpleural areas, and resolution of pleural effusions, indicating a positive response to treatment; 3, Axial chest CT images showed a decrease in density and a slight reduction in the extent of bilateral ground-glass opacities; 4, Contrast-enhanced axial chest CT images performed during the second hospitalization showed extensive ground-glass opacities with significant densification in both lungs involving more than 75% of lung volume, and the appearance of a 14 mm thick right pleural effusion; 5, Axial chest CT images performed during the third hospitalization showed multiple bilateral ground-glass opacities, with areas showing hyperdense opacities and condensation with a retractile appearance, accompanied by traction bronchiectasis; 6, Axial chest CT images showed a slight intensification of bilateral ground-glass opacities, regression of consolidations in the upper lobes with increased bronchiectasis features in the lower lobes, alongside near-complete regression of pleural effusions, suggesting a discordant evolution of interstitial involvement with potential lower lobar infection; 7, Axial chest CT images showed a worsening of bilateral panlobar pneumonia with increased consolidation volume and size of confluence foci, alongside the emergence of a mild bilateral pleural effusion; 8, Axial chest CT images performed after the third hospitalization showed a decrease in bilateral ground-glass opacities and a reduction of consolidative opacities in the lower lobes, right upper lobe, and lingula, with less prominent bronchiectasis features.

**Supplementary Figure S2.**
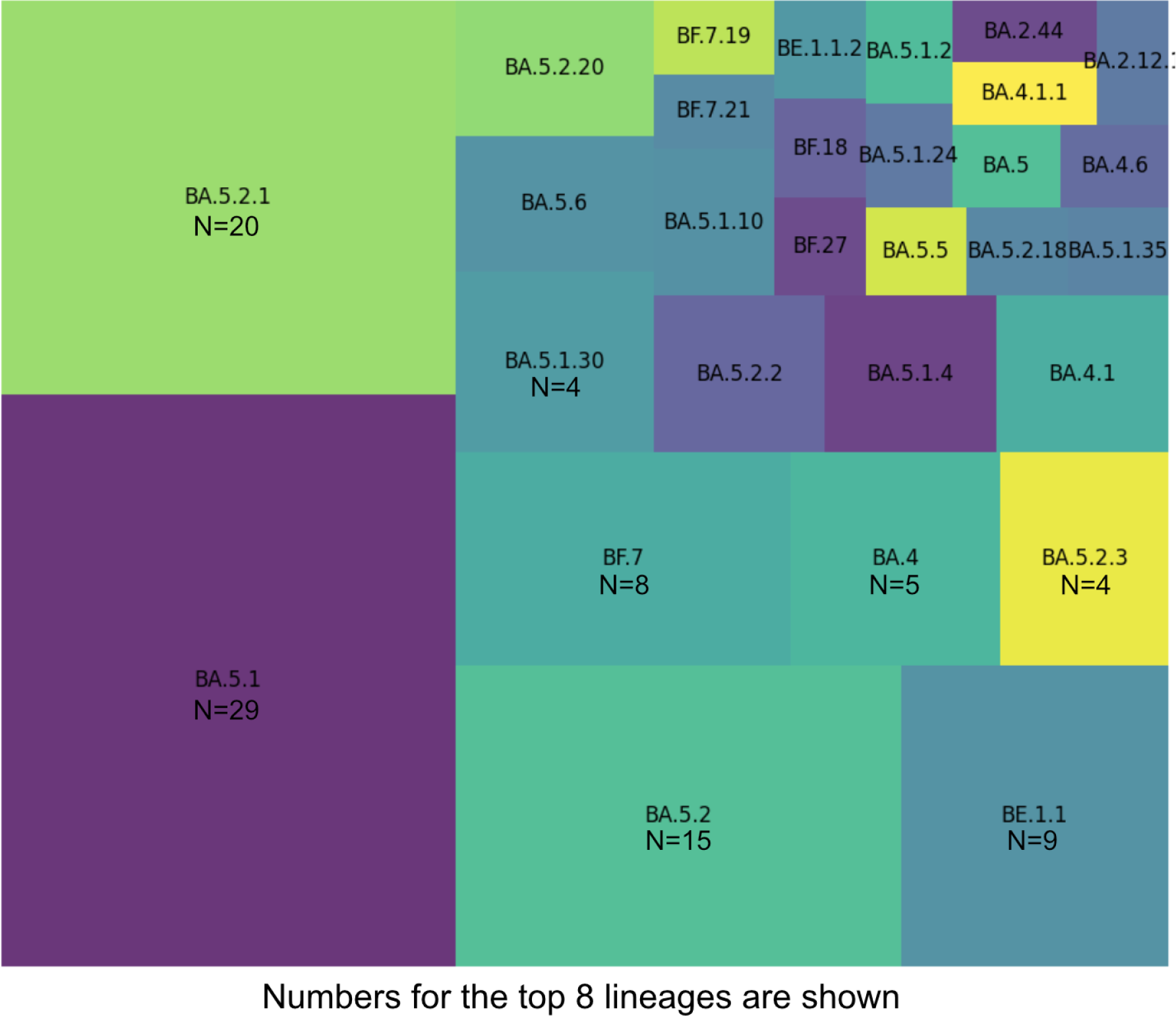
Lineages observed in Liège, Belgium during the week of the patients 1^st^ hospitalisation for COVID-19 pneumopathy, July 2022.

**Supplementary Figure S3.**
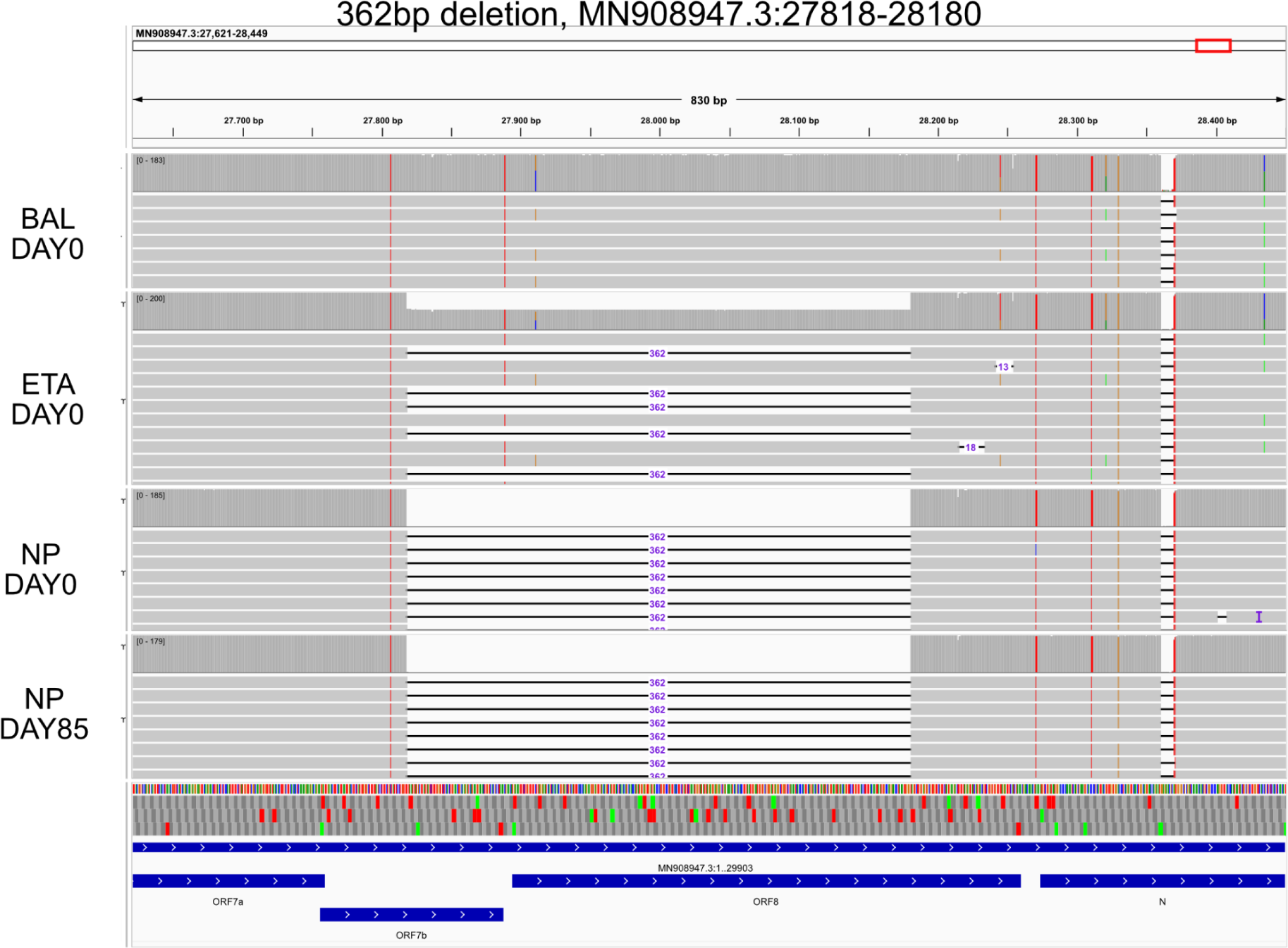
Long range PCR and nanopore sequencing of the 362bp deletion impacting ORF7b and ORF8 observed in the NP sample and a subset of the reads in the ETA sample.

**Supplementary Figure S4.**
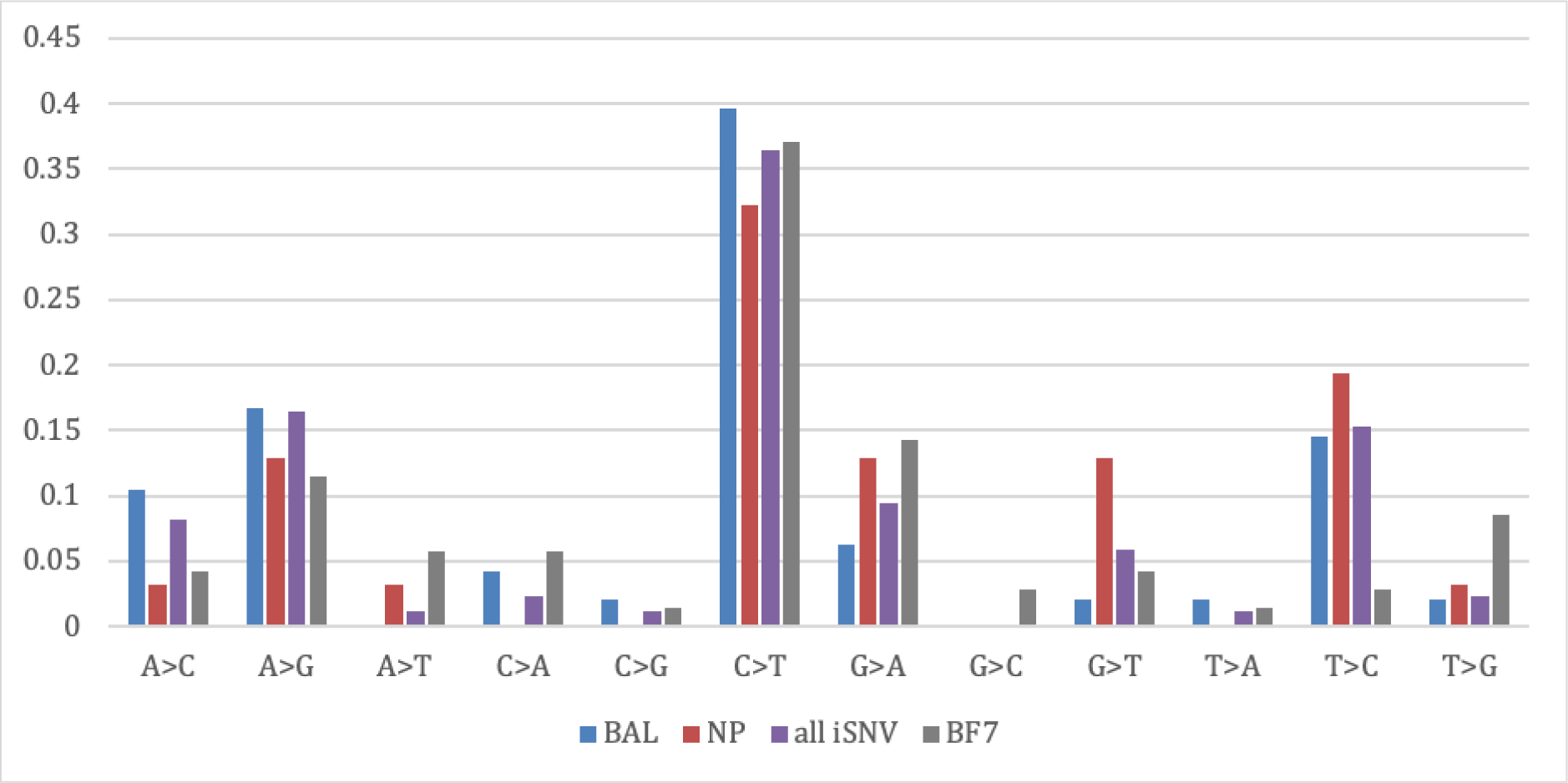
Mutation spectra of iSNVs in BAL, NP and all iSNVs, as well as SNVs in ancestral BF.7 lineage.

**Supplementary Figure S5.**
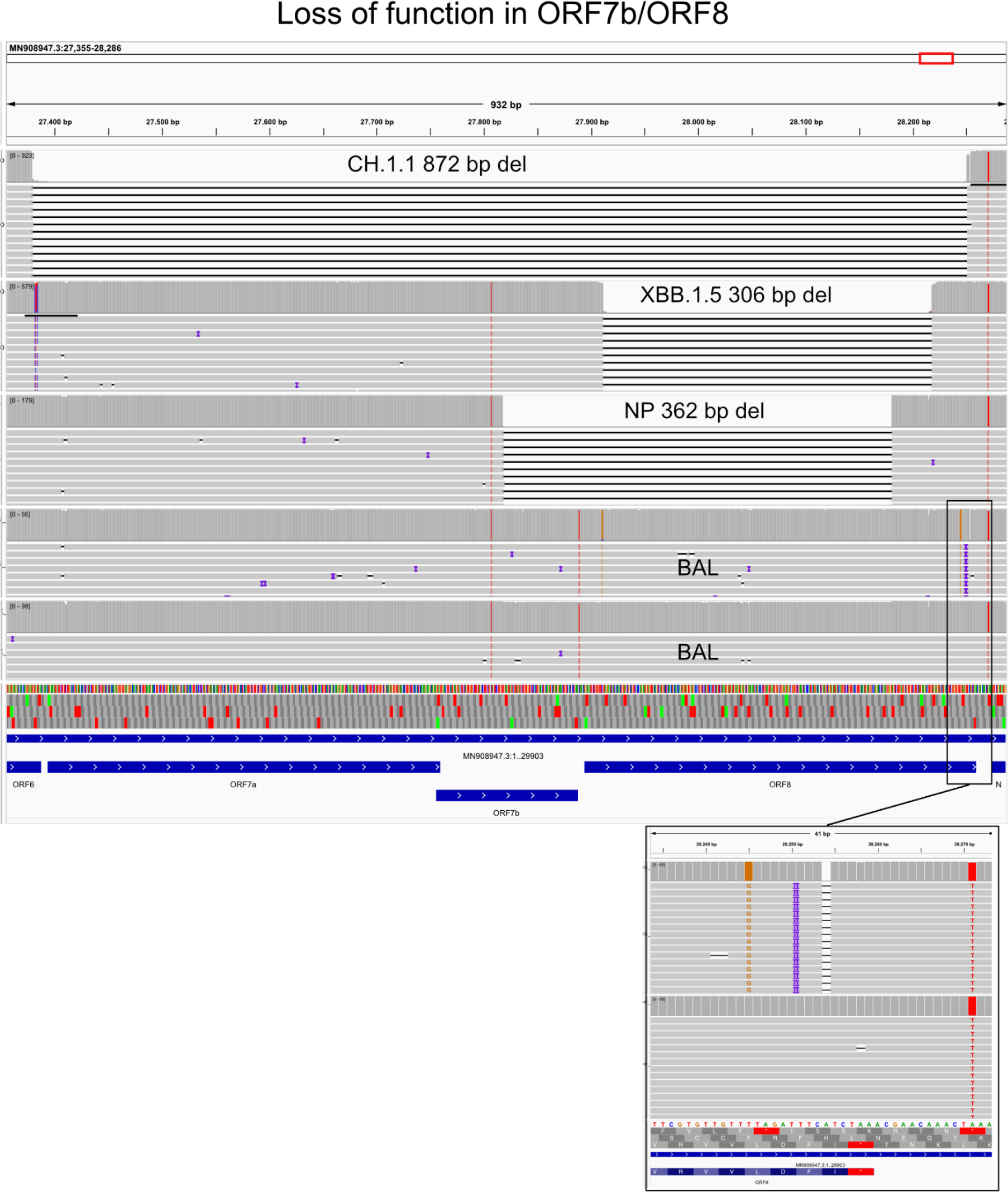
Long-range PCR across to ORF7a/b/8 sequenced with nanopore. Two examples of large deletions in circulating lineages (CH.1.1 and XBB.1.5) were observed during routine SARS-CoV-2 genomic surveillance at the University of Liège (Belgium) in 2023. The deletion seen in the NP sample is shown below these. The last two panels show the reads from the BAL separated based on whether they carry the ORF8 frameshift or the wild-type sequence.

**Supplementary Figure S6.**
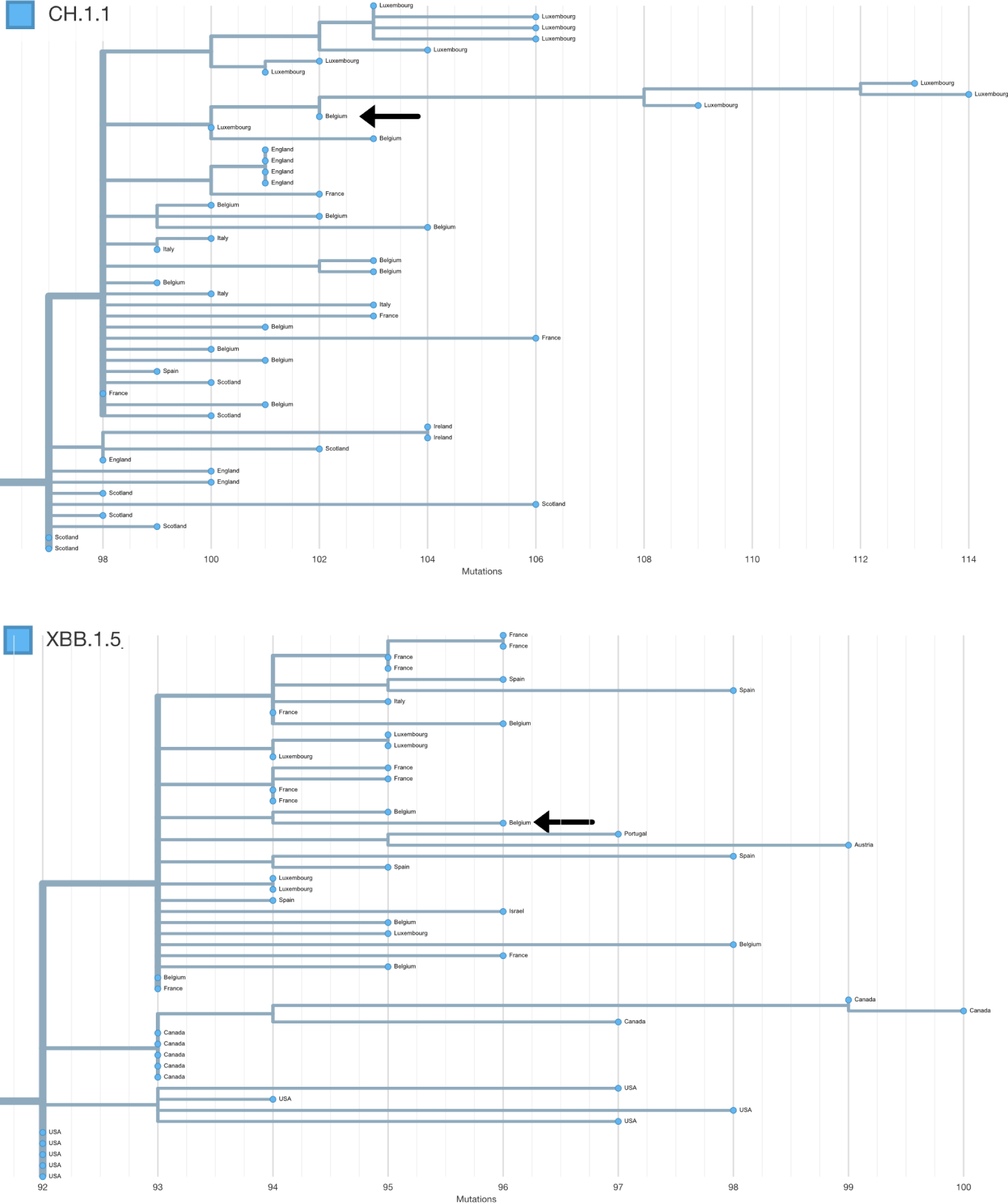
Neighbouring samples in the global phylogenetic tree generated by UShER for the CH.1.1 sample with ORF7a/b/8 deletion and the XBB.1.5. sample with ORF8 deletion. Both are indicated by an arrow.

**Supplementary Table S2.** Estimated impact of spike amino acid changes, values taken from Dadonaite *et al*. (2023)^19^. GISAID search was performed 2024-01-18, Low coverage genomes were excluded. Abbreviation: BAL = bronchoalveolar, NP = nasopharyngeal

| AA change | Site | Number on GISAID | Max allele frequency | XBB AA | mut AA | Human sera escape | ACE2 binding | S region |
| --- | --- | --- | --- | --- | --- | --- | --- | --- |
| S:I19L | BAL | 6354 | 0.22 | I | L | -0.06043 | 0.1969 | NTD |
| S:T124I | BAL | 782 | 0.54 | T | I | -0.01891 | -0.1663 | NTD |
| S:T274I | BAL | 1774 | 0.24 | T | I | 0.04957 | -0.01831 | NTD |
| S:P330S | BAL | 3432 | 0.95 | P | S | -0.162 | 0.2282 | other |
| S:F375S | BAL | 8999* | 0.99 | F | S | -0.6879 | 0.8739 | RBD |
| S:E619A | BAL | 175 | 0.98 | E | A | NaN | 0.8979 | other |
| S:H146Y# | NP | 7305 | 0.88 | Q | Y | 0.11 | -0.09042 | NTD |
| S:T523I | NP | 147 | 1 | T | I | -0.5882 | 1.253 | RBD |
| S:V551I | NP | 1521 | 0.98 | V | I | NaN | -0.153 | other |
| S:V976A | NP | 112 | 0.99 | V | A | 0.4773 | -0.3908 | S2 |
| S:N1134H | NP | 20 | 0.98 | N | H | -0.00334 | -0.1161 | S2 |
| S:G1219V# | NP | 16166 | 0.94 | G | V | -0.001387 | -0.2215 | other |

\*As this was a reversion, in addition to searching for samples that did not have S375F (-Spike\_S375F) also searched for the adjacent mutation A22688G to select for Omicron genomes and remove samples with gaps in region

## Notes

### Competing Interest Statement

The authors have declared no competing interest.

### Funding Statement

This work was supported in part by the Region Wallone project WALGEMED (convention no. 1710180) and the FNRS (H.C.008.20). Sequencing was done as a part of the National Genomic Surveillance Platform for SARSCoV2 in Belgium. SD acknowledges support from the Fonds National de la Recherche Scientifique (F.R.S. FNRS, Belgium; grant F.4515.22), from the Research Foundation Flanders (Fonds voor Wetenschappelijk Onderzoek Vlaanderen, FWO, Belgium; grant G098321N), and from the European Union Horizon 2020 projects MOOD (grant agreement 874850) and LEAPS (grant agreement 101094685).

### Author Declarations

The Ethics committee/IRB of the University Hospital Liege gave ethical approval for this work. Reference number: 2020-139.

